# MPRAGE-derived quantitative T_1_ mapping to assess diffuse white matter alterations in multiple sclerosis

**DOI:** 10.64898/2026.05.04.26351019

**Authors:** Audrey Lavielle, Fanny Munsch, Aurélie Ruet, Thomas Tourdias, Yannick Crémillieux

**Author notes:** **Corresponding author**: Yannick Crémillieux, Institut des Sciences Moléculaires, CNRS, Université de Bordeaux, 33076 Bordeaux, France.

## Abstract

**Background:** Multiple sclerosis (MS) is characterized by focal white matter (WM) lesions, but subtle damage also occurs in normal-appearing white matter (NAWM). We developed a method to generate quantitative T_1_ maps from MPRAGE (Magnetization Prepared Rapid Gradient Echo) images and evaluated its ability to detect NAWM abnormalities across different MS phenotypes.

**Methods:** T_1_ maps were derived from MPRAGE using a theoretical signal model and compared with MP2RAGE (Magnetization Prepared 2 Rapid Gradient Echoes) T_1_ values in four healthy volunteers. The method was then applied to 87 MS patients, divided into clinically isolated syndrome (CIS), relapsing–remitting MS (RRMS), and primary progressive MS (PPMS), with age- and sex-matched healthy controls. T_1_ was measured in NAWM and lesions. Histogram analysis provided mean T_1_, full width at half maximum (FWHM), and skewness.

**Results:** In healthy volunteers, T_1_ values matched MP2RAGE. In controls matched to the MS cohort, T_1_ increased with age (r = 0.35, p < 0.05). CIS patients showed no significant differences in any metric. RRMS and PPMS patients showed unchanged mean NAWM T_1_ but significantly different distributions, with higher FWHM (p<0.05) and skewness (p<0.001). An increase in T_1_ values was observed in MS lesions compared to NAWM in all groups.

**Conclusion:** This study confirms the feasibility of deriving quantitative T_1_ maps from standard MPRAGE, offering reliable information to facilitate MS monitoring without additional acquisitions.

## 1. Introduction

Multiple sclerosis (MS) is characterized by the presence of focal lesions corresponding to areas of demyelination, which can be detected using conventional magnetic resonance imaging (MRI). However, lesion volume, commonly referred to as lesion load, only moderately correlates with clinical disability, due to the presence of pathological changes extending beyond MRI-visible lesions (Filippi et al., 2019). Histopathological studies have demonstrated that demyelination, inflammation, edema, axonal damage, and gliosis are not confined to focal lesions but also affect Normal-Appearing White Matter (NAWM) (Kornek et al., 2000). Conventional MRI sequences, such as Fluid Attenuated Inversion Recovery (FLAIR) or T_1_-and T_2_-weighted imaging, are unable to identify these subtle and diffuse alterations within the NAWM.

In contrast, advanced MRI techniques have proven to be more sensitive to microstructural and biochemical changes in MS and have been shown to provide clinically relevant insights into disease progression. Semi-quantitative approaches, such as magnetization transfer imaging (Filippi et al., 2000; Khaleeli et al., 2007; Tortorella et al., 2000) and diffusion tensor imaging (Bammer et al., 2000; Filippi et al., 2001), have revealed abnormalities in NAWM and Normal-Appearing Gray Matter (NAGM) that are undetectable by conventional MRI.

In parallel, quantitative methods such as T_1_ relaxation time mapping provide objective metrics of tissue damage and have proven particularly valuable for characterizing pathological changes in both focal lesions and NAWM. Several studies have reported increased T_1_ values in NAWM in MS patients compared to healthy controls (HC) (Bluestein et al., 2012; Gracien et al., 2016a; Parry et al., 2002; Srinivasan et al., 2003; Steenwijk et al., 2016; Vaithianathar et al., 2002; Vrenken et al., 2006). T_1_ relaxation time is influenced by the amount of free water in tissue and is therefore sensitive to pathological processes such as edema, inflammation, gliosis, and axonal loss, all of which contribute to increased extracellular space and increased T_1_ values. Several MRI sequences have been proposed for in vivo T_1_ mapping. Inversion and saturation recovery methods are considered reference standards but remain impractical in vivo due to long acquisition times (Crawley and Henkelman, 1988). Faster alternatives include Look-Locker–based and variable flip angle (VFA) techniques (Look and Locker, 1970; Stikov et al., 2015), although the latter is sensitive to B_1_ inhomogeneities (Subashi et al., 2014). More recently, the Magnetization Prepared 2 Rapid Gradient Echo (MP2RAGE) sequence, an extension of the Magnetization Prepared RApid Gradient Echo (MPRAGE) sequence, has been introduced as a robust and rapid 3D T_1_ mapping approach with reduced B_1_ sensitivity (Marques et al., 2010). Despite its demonstrated advantages (Kober et al., 2012; Simioni et al., 2014) – particularly its superior performance in T_1_ mapping with only a modest increase in acquisition time – the MP2RAGE sequence remains rarely incorporated in routine clinical practice. This limited adoption likely stems from either persistent barrier to clinical acceptance or restricted sequence availability in hospital radiology departments.

In contrast, the vast majority of clinical protocols for patients with neurodegenerative diseases, such as MS, routinely include the T_1_-weighted MPRAGE sequence, widely recognized as a reference imaging sequence in neuroradiology for MS patients (Brisset et al., 2020). This extensive use of MPRAGE in neuroradiology protocols has led, for example, to recent developments involving machine learning-based approaches for obtaining quantitative T_1_ maps from standard MPRAGE and FLAIR acquisitions (Snyder et al., 2025).

Given this context, and considering the extensive use of MPRAGE in both past and ongoing MRI studies of MS patients, we investigated the feasibility and clinical relevance of a previously reported method for generating quantitative T_1_ maps from MPRAGE acquisitions (Lavielle et al., 2023). Specifically, our study aimed to assess the ability of this approach to detect subtle abnormalities in normal-appearing white matter (NAWM) across different MS phenotypes.

## 2. Materials and methods

### 2.1. Participants

Initially, four healthy volunteers were enrolled in this study to provide a preliminary assessment of the proposed approach.

The approach was then extended to a cohort of MS patients and matched HC. We prospectively included 87 patients with MS and age- and sex-matched HCs from three independent studies: MICROSEP, AUBACOG, and PROCOG (clinical trial registration numbers: NCT03692975, NCT03768648, and NCT03455582, respectively). These studies were primarily designed to investigate the underlying mechanisms of cognitive impairment across different stages of MS. Participants in the MICROSEP cohort were diagnosed with Clinically Isolated Syndrome (CIS) or early MS. Inclusion occurred within six months following a first clinical episode suggestive of an inflammatory demyelinating event, regardless of its clinical presentation. Eligible patients were required to have at least two clinically silent lesions ≥ 3 mm on T_2_-weighted brain or spinal MRI, including at least one cerebral lesion that was ovoid or periventricular. Patients were classified as having early MS if they met the 2017 McDonald criteria for dissemination in time and space, and as CIS otherwise. Participants from the AUBACOG and PROCOG studies were diagnosed with Relapsing-Remitting MS (RRMS) and Primary Progressive MS (PPMS), respectively, according to the 2017 McDonald criteria.

For each subgroup, age- and sex-matched HC were included: 43 HC for the CIS group, 35 for the RRMS group, and 26 for the PPMS group. All participants provided written informed consent prior to enrollment and the study was approved by the local ethics committee.

The study was sponsored by the University of Bordeaux is referenced as RCB 2016-A00434- 47. MICROSEP (NCT03692975), AUBACOG (NCT03768648), and PROCOG (NCT03455582) have been approved by the institutional review boards under the following approval numbers: MICROSEP (#18.021, Comité de Protection des Personnes Ile de France V), AUBACOG (#18 78, Comité de Protection des Personnes Sud Méditerranée I), PROCOG (#2017-72, Comité de Protection des Personnes Nord Ouest II).

### 2.2. MRI protocol

MRI scans were performed using a 3 T Vantage Galan ZGO scanner (Canon Medical Systems Corporation, Tokyo, Japan) with a whole-body coil for RF excitation, and a 32-channel head coil for reception.

For the four healthy volunteers, T_1_-weighted images were acquired using a 3D-MPRAGE sequence with the following parameters: echo time: 2.8 ms; repetition time: 2500 ms; inversion time: 950 ms, flip angle: 9°, image size: 180 x 224 x 224; field of view: 180 x 224 x 224 mm^3^; spatial resolution: 1 x 1 x 1 mm^3^; total acquisition time: 6 min 18 s. A bias field correction related to radiofrequency inhomogeneities was applied using the FAST tool from the FMRIB Software Library (version 6.0.6; FMRIB, Oxford, UK) (Jenkinson et al., 2012; Smith et al., 2004). In addition, T_1_ maps were generated from an MP2RAGE acquisition (Marques et al., 2010), which provides a well-validated method for quantitative T_1_ mapping. The sequence parameters were: echo time: 3.3 ms; repetition time: 7.5 ms; inversion times: 655/3300 ms; image size: 384 x 384 x 50; field of view: 190 x 190 x 150 mm^3^; spatial resolution: 0.5 x 0.5 x 3.0 mm^3^; total acquisition time: 7 min 42 s. For the MS cohort (patients and matched controls), the imaging protocol included both a 3D-MPRAGE acquisition with the same parameters as above and a 3D-FLAIR sequence to identify MS lesions. The 3D-FLAIR images were acquired with the following parameters: echo time: 444.5 ms; repetition time: 7000 ms; inversion time: 2100 ms; image size: 180 x 224 x 224; field of view: 180 x 224 x 224 m^3^; spatial resolution: 1 x 1 x 1 mm^3^; total acquisition time: 4 min 54 s.

### 2.3. Image segmentation

The segmentation of MPRAGE images was performed using AssemblyNet software (Coupé et al., 2020), available through the volBrain platform (Manjon and Coupe, 2016). AssemblyNet is based on a set of 250 deep learning models that allow for the fully automated segmentation of the brain into 132 anatomical structures. In this study, it was used to segment the white matter (WM), gray matter (GM), and cerebrospinal fluid (CSF). The cingulate cortex was also specifically extracted (see below).

MS lesions were segmented manually based on the 3D-FLAIR images by an experienced neuroradiologist, based on the identification of typical hyperintense signals associated with MS. The NAWM was then defined as the portion of WM that excludes lesions, obtained by subtracting the MS lesion masks from the total WM segmentation.

### 2.4. Computation of T_1_ maps from MPRAGE acquisitions

The T_1_ maps were obtained from MPRAGE acquisitions using a theoretical approach described in a previous study (Lavielle et al., 2023). Briefly, this approach relies on an analytical expression of the MPRAGE signal amplitude (Gowland and Leach, 1992), which depends on sequence parameters and tissue properties such as T_1_, T_2_*, and proton density. In practice, the signal intensity in a region with a known T_1_ value (either a reference tissue or an external reference) is used to scale the relationship between signal intensity and T_1_ value. Variations in proton density across tissues (WM, GM, CSF) are then considered so that the signal intensity of each voxel can be converted into a quantitative T_1_ value. In this way, a 3D T_1_ map can be generated from a MPRAGE acquisition (Lavielle et al., 2023).

In this study, the cingulate cortex was chosen as the reference tissue because of its stability across individuals and the minimal differences observed between MS patients and HC (Steenwijk et al., 2016). Its T_1_ value was fixed to 1.369 s, in agreement with the mean T_1_ measured using the MP2RAGE method and consistent with values reported in the literature (Bojorquez et al., 2017; Hagiwara et al., 2021; Steenwijk et al., 2016).

For the computation of T_1_ maps, the proton density values for WM, GM and CSF were set to 66%, 83% and 100% of the proton density of pure water at 37°C for healthy subjects, and to 69%, 80% and 100% for MS patients, respectively, based on values reported in the literature (Gracien et al., 2016b; Hagiwara et al., 2021).

### 2.5. Analysis of NAWM

The distribution of T_1_ values within the NAWM was analyzed for each subject. In addition, for each MS phenotype, a global histogram was generated by pooling the NAWM voxels of all controls and, separately, of all patients within the group. To characterize the T_1_ distribution, each histogram was constructed with a bin size of 0.5 milliseconds, normalized by the number of voxels and modeled as the sum of Gaussian functions.

The optimal number of Gaussian components was determined using the Bayesian Information Criterion (BIC), a metric that evaluates the goodness of fit while penalizing model complexity to avoid overfitting. In all cases, the BIC systematically selected a two-Gaussian model.

The histograms were characterized using key metrics, including the mean, standard deviation and full width at half maximum (FWHM). Additionally, the skewness, defined as the adjusted Fisher-Pearson standardized moment coefficient, was computed to quantify the asymmetry of the distribution. Skewness measures the symmetry of a distribution relative to its mean and reflects its tendency to stretch further to the right or left. A positive skewness indicates a longer or larger right histogram tail compared to the left, suggesting that a subset of voxels exhibits higher T_1_ values, whereas a negative skewness reflects the opposite trend. This analysis provides insights into potential deviations in T_1_ distribution within NAWM.

All histogram construction and analyses were performed using MATLAB R2024b (MathWorks, Natick, MA).

### 2.6. Statistical analysis

All statistical analyses were performed using RStudio (PBC, Boston, MA), and outliers were excluded using the interquartile range method. Shapiro-Wilk tests were used to assess the normality of the variables (O’Shaughnessy et al., 2025). When data followed a normal distribution, parametric tests were applied, including Student’s t-test for comparisons between two groups and analysis of variance (ANOVA) for comparisons across multiple groups. When the assumption of normality was rejected, non-parametric alternatives were used, specifically the Mann–Whitney U test for two-group comparisons and the Kruskal–Wallis test for comparisons across more than two groups.

Additionally, correlation analyses were conducted using Pearson’s correlation test to explore relationships between variables following a normal distribution, such as age, lesion volume, and histogram-derived parameters (e.g., mean T_1_ values, FWHM and skewness). A significance threshold of p < 0.05 was applied to determine statistical significance.

Finally, a second-degree polynomial fit was used to model the relationship between T_1_ values in the WM and age for all HC.

## 3. Results

### 3.1. Demographic and clinical characteristics of the study cohort

The demographic and clinical characteristics of the study cohort are summarized in Table 1. Age and sex distribution were similar between MS patients and healthy controls within each phenotype group.

**Table 1.** Demographic and clinical characteristics. CIS indicates Clinically Isolated Syndrome; RRMS indicates Relapsing-Remitting Multiple Sclerosis; PPMS indicates Primary Progressive Multiple Sclerosis; EDSS indicates Expanded Disability Status Scale; NAWM indicates Normal-Appearing White Matter.

| **Group type** | **CIS**<br>**(n=32)** | **Matched-**<br>**CIS-**<br>**controls**<br>**(n=43)** | **RRMS**<br>**(n=30)** | **Matched-**<br>**RRMS-**<br>**controls**<br>**(n=35)** | **PPMS**<br>**(n=25)** | **Matched-**<br>**PPMS-**<br>**controls**<br>**(n=26)** |
| --- | --- | --- | --- | --- | --- | --- |
| **Age (years)** | 35 ± 10 | 33 ± 7 | 43 ± 10 | 41 ± 6 | 52 ± 7 | 51 ± 6 |
| **Female/male** | 25/7 | 35/8 | 19/11 | 21/14 | 15/10 | 20/6 |
| **EDSS score** | 1,7 ± 1,1 | - | 1,4 ± 1,2 |  | 3,6 ± 1,3 |  |
| **Disease**<br>**duration**<br>**(months)** | 0,4 ± 0,1 | - | 9,9 ± 7,0 |  | 4,4 ± 3,1 |  |
| **NAWM**<br>**volume (mL)** | 426 ± 45 | 427 ± 52 | 416 ± 51 | 451 ± 48 | 407 ± 84 | 420 ± 44 |
| **Lesion**<br>**volume (mL)** | 3.0 ± 2.1 | - | 8.1 ± 6.4 | - | 15.8 ±<br>26.1 | - |

**Table 2.** Mean and standard deviation of T_1_ values for 4 volunteers obtained from MPRAGE and MP2RAGE acquisitions in the following brain regions: occipital and frontal WM, frontal and occipital GM, putamen, caudate nucleus and CSF. MPRAGE indicates Magnetization Prepared RApid Gradient Echo; MP2RAGE indicates Magnetization Prepared 2 RApid Gradient Echo; WM indicates White Matter; GM indicates Gray Matter; CSF indicates CerebroSpinal Fluid.

|  | **MPRAGE-derived**<br>**$T_1$ (s)** | **$T_1$ literature value (s)**<br>**(Bojorquez et al., 2017;**<br>**Hagiwara et al., 2021)** | **MP2RAGE-derived $T_1$**<br>**(s)** |
| --- | --- | --- | --- |
| **Occipital WM** | $0.958 \pm 0.016$ | 0.700 – 1.000 | $0.958 \pm 0.082$ |
| **Frontal WM** | $0.978 \pm 0.058$ | | $0.966 \pm 0.093$ |
| **Occipital GM** | $1.301 \pm 0.059$ | 1.000 – 1.600 | $1.403 \pm 0.150$ |
| **Frontal GM** | $1.396 \pm 0.055$ | | $1.440 \pm 0.170$ |
| **Putamen** | $1.203 \pm 0.061$ | 1.100 – 1.300 | $1.207 \pm 0.042$ |
| **Caudate**<br>**Nucleus** | $1.250 \pm 0.022$ | 1.200 – 1.400 | $1.249 \pm 0.064$ |
| **CSF** | $2.092 \pm 0.195$ | 3.800 – 5.000 | $2.792 \pm 1.499$ |

### 3.2. Evaluation on healthy volunteers

Fig. 1.A shows an example of a T_1_ map generated from an MPRAGE acquisition in a healthy volunteer using the proposed approach. The spatial resolution of the T_1_ map is the same than the MPRAGE image, namely 1 x 1 x 1 mm^3^. For comparison, Fig.1.B presents the T_1_ map obtained from the MP2RAGE acquisition, with a spatial resolution of 0.5 x 0.5 x 3.0 mm^3^. T_1_ measurements in several brain regions were performed in the four healthy volunteers, and the corresponding mean ± standard deviations are summarized in Table . The mean MPRAGE-derived T_1_ values measured in brain tissues were found in the range of values reported in the literature (Bojorquez et al., 2017; Hagiwara et al., 2021) and similar to those obtained with the MP2RAGE sequence. These results are supported by the Bland-Altman plot in Fig. 2, which shows the difference in T_1_ values obtained from MPRAGE and MP2RAGE acquisitions for each brain region. The small bias and narrow limits of agreement indicate a good overall agreement between the two methods. The T_1_ values measured in the CSF with the two approaches differ greatly from the values reported in the literature.

**Fig. 1.**
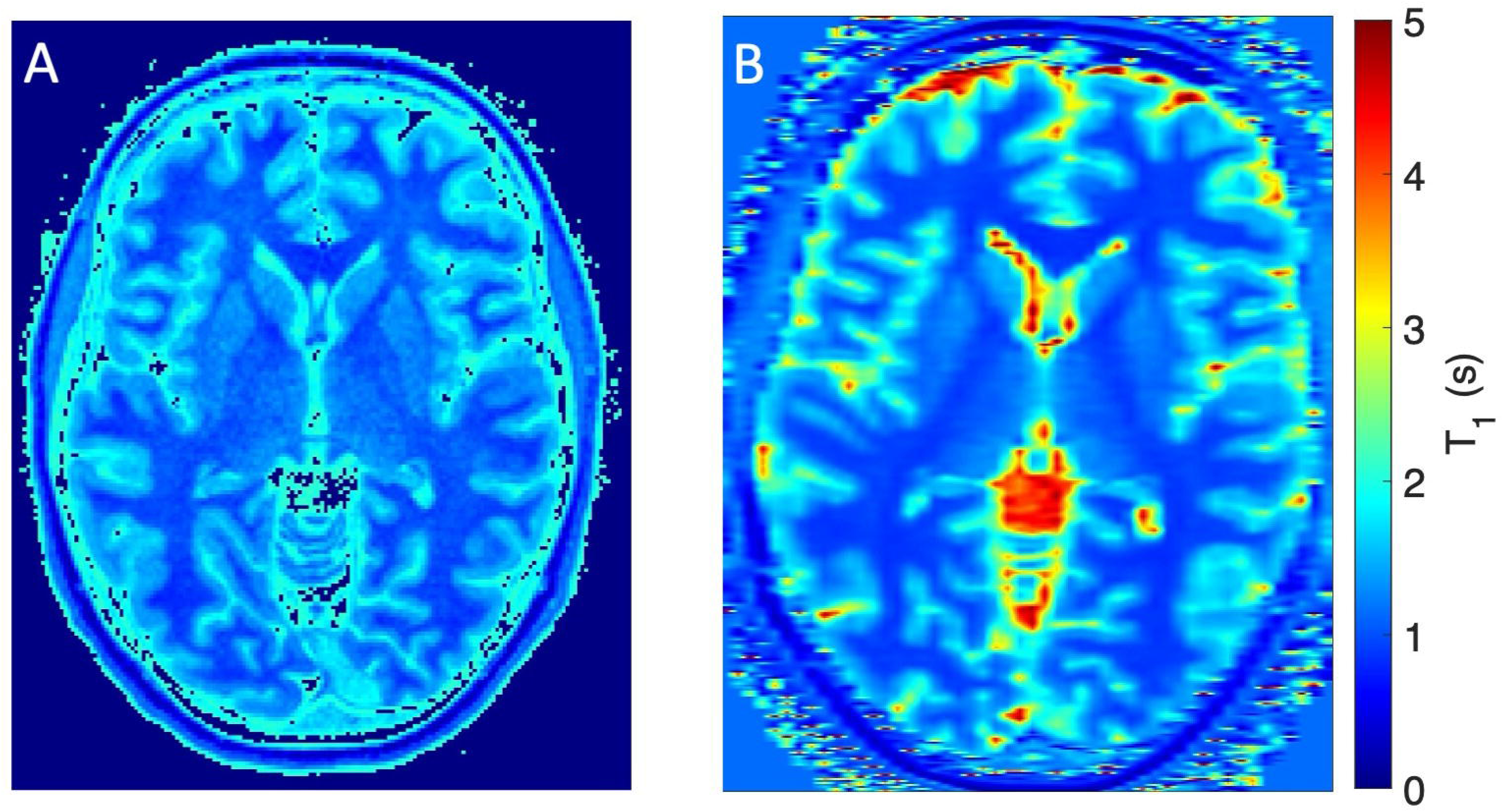
Comparison of T_1_ maps obtained from MPRAGE and MP2RAGE sequences in a healthy volunteer. (A) T_1_ map derived from a MPRAGE image acquired from a healthy volunteer. (B) MP2RAGE T_1_ map obtained at the same position. MPRAGE indicates Magnetization Prepared RApid Gradient Echo; MP2RAGE indicates Magnetization Prepared 2 RApid Gradient Echo.

**Fig. 2.**
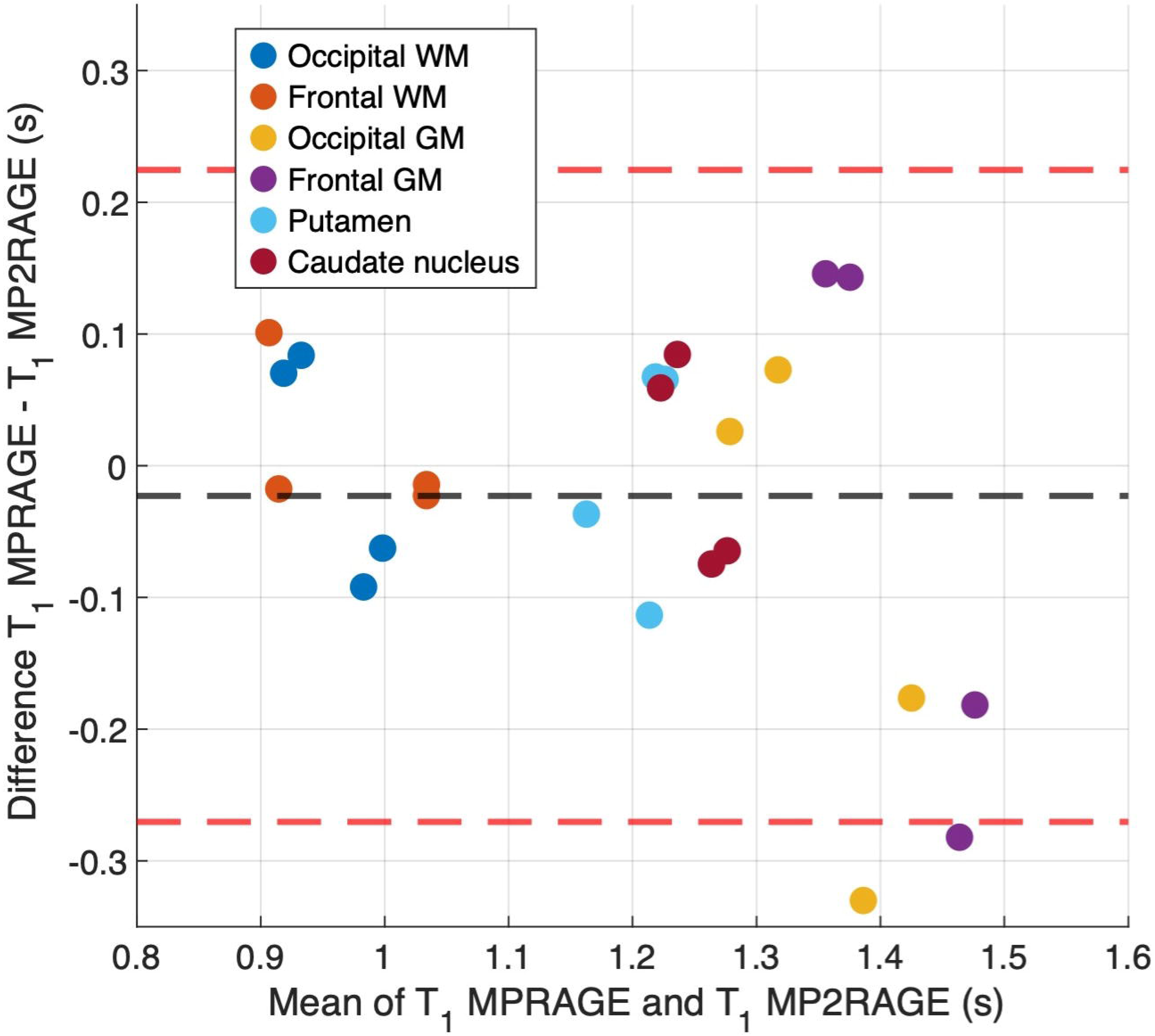
Bland-Altman plot comparing T_1_ values measured from MPRAGE and MP2RAGE acquisitions. T_1_ values were measured in four healthy volunteers across the following brain regions: occipital and frontal WM, occipital and frontal GM, putamen and caudate nucleus. The dashed black line represents the mean difference, and the red dashed lines indicate 95% limits of agreement. MPRAGE indicates Magnetization Prepared RApid Gradient Echo; MP2RAGE indicates Magnetization Prepared 2 RApid Gradient Echo; WM indicates White Matter; GM indicates Gray Matter.

### 3.3. Application to the MS cohort

#### 3.2.1. Computation of T_1_ maps

The computation of a T_1_ map from the MPRAGE acquisition is illustrated in Fig. 3, which shows FLAIR, MPRAGE, and the corresponding T_1_ maps in a PPMS patient and in her matched HC. White and pink contours delineate the NAWM (or WM in the HC) and MS lesions, respectively, used for T_1_ measurements. Once the preprocessing steps (segmentation of the different tissues and lesions) were completed, the computation of a 3D T_1_ map from a MPRAGE acquisition required approximately one minute on a standard computer. The process was stable across subjects, and no issues were encountered in the generation of the maps.

**Fig. 3.**
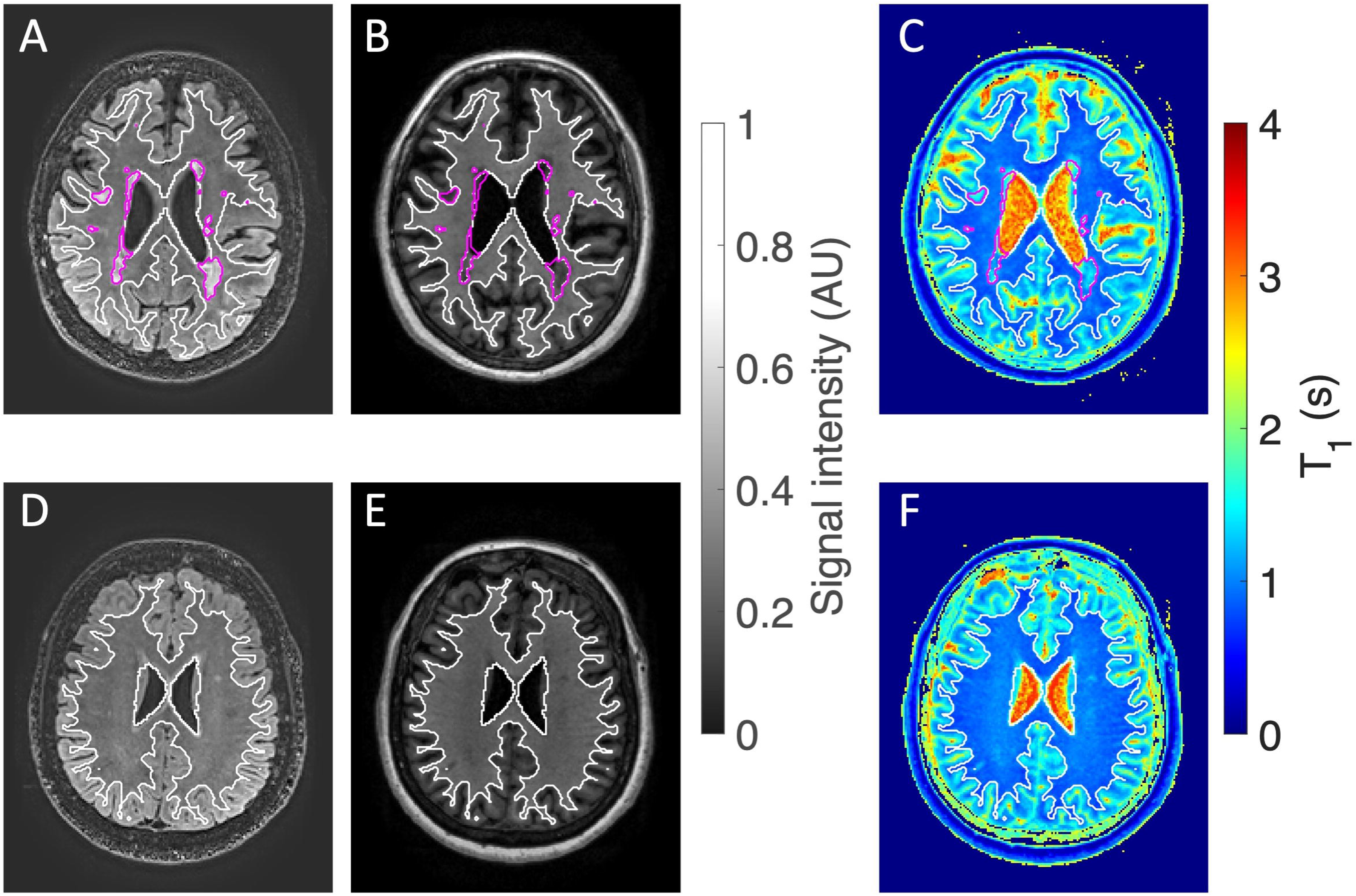
FLAIR, MPRAGE, and T_1_ maps derived from MPRAGE images obtained in a patient with PPMS (top row) and her matched HC (bottom row). (A, D) FLAIR images. (B, E) MPRAGE images. (C, F) T_1_ maps derived from the MPRAGE images. White contours delineate NAWM in the patient and WM in the HC, while pink contours indicate lesions in the patient. HC indicates Healthy Control; MPRAGE indicates Magnetization Prepared RApid Gradient Echo; FLAIR indicates Fluid Attenuated Inversion Recovery; PPMS indicates Primary Progressive Multiple Sclerosis; NAWM indicates Normal-Appearing White Matter; WM indicates White Matter.

#### 3.2.2. T_1_ measurements in WM of healthy controls

An age-related increase in WM T_1_ values was observed in HC (Fig. 4.A). Specifically, the mean T_1_ values were 0.885 ± 0.019 s in the CIS-matched control group (33 ± 7 years), 0.884 ± 0.016 s in the RRMS-matched control group (41 ± 6 years), and 0.894 ± 0.019 s in the PPMS-matched control group (51 ± 6 years). However, these differences between groups were not statistically significant. Nevertheless, a weak but statistically significant positive correlation was observed between mean T_1_ values and age (r = 0.35; p < 0.05). In addition, a quadratic model was fitted to the data, as illustrated in Fig. 4.A, according to the equation: *T*_1_ = 2.8 ∗ 10^−5^ ∗ *Age*^2^ + 1.6 ∗ 10^"%^ ∗ *Age* + 0.9. This regression yielded a coefficient of determination *R*^2^ = 0.16, illustrating a general trend of increasing T_1_ values with age across all subjects. In contrast, no significant correlation was found between age and the FWHM (r = 0.06; p = 0.52), nor with histogram skewness (r = 0.2; p = 0.06), suggesting that the shape of the T_1_ distribution remains globally stable with age, as illustrated in Fig. 4.B.

**Fig. 4.**
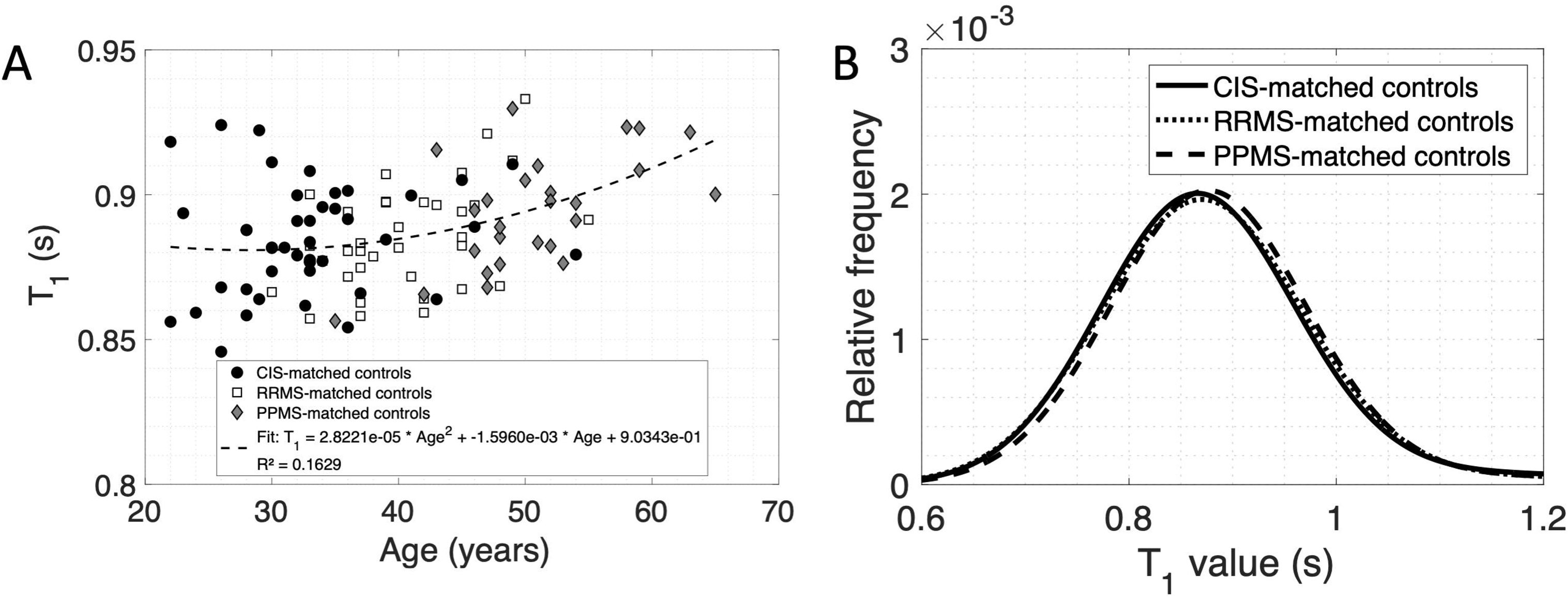
T_1_ measurements in the WM of healthy controls. (A) Variation of T_1_ as a function of age. (B) Distribution of T_1_ values across different control groups. CIS indicates Clinically Isolated Syndrome; RRMS indicates Relapsing-Remitting Multiple Sclerosis; PPMS indicates Primary Progressive Multiple Sclerosis; WM indicates White Matter.

#### 3.2.3. T_1_ measurements in the NAWM and lesions of MS patients

Fig. 5A presents the distributions of T_1_ values in the NAWM of MS patients across different phenotypes. The mean T_1_ values, FWHM, and skewness measurements in the NAWM of MS patients and the corresponding WM of HC are summarized in Table 2. For each group, the reported values correspond to the mean and standard deviation calculated from individual subject measurements. Fig. 6. illustrates these results using boxplots comparing the mean T_1_ values, FWHM, and skewness between MS patients and their matched controls. Notably, all measured skewness values were positive, indicating that the asymmetry observed in the distributions was due to a longer or larger right tail. This suggests that a subset of voxels exhibited higher T_1_ values across all groups.

**Fig. 5.**
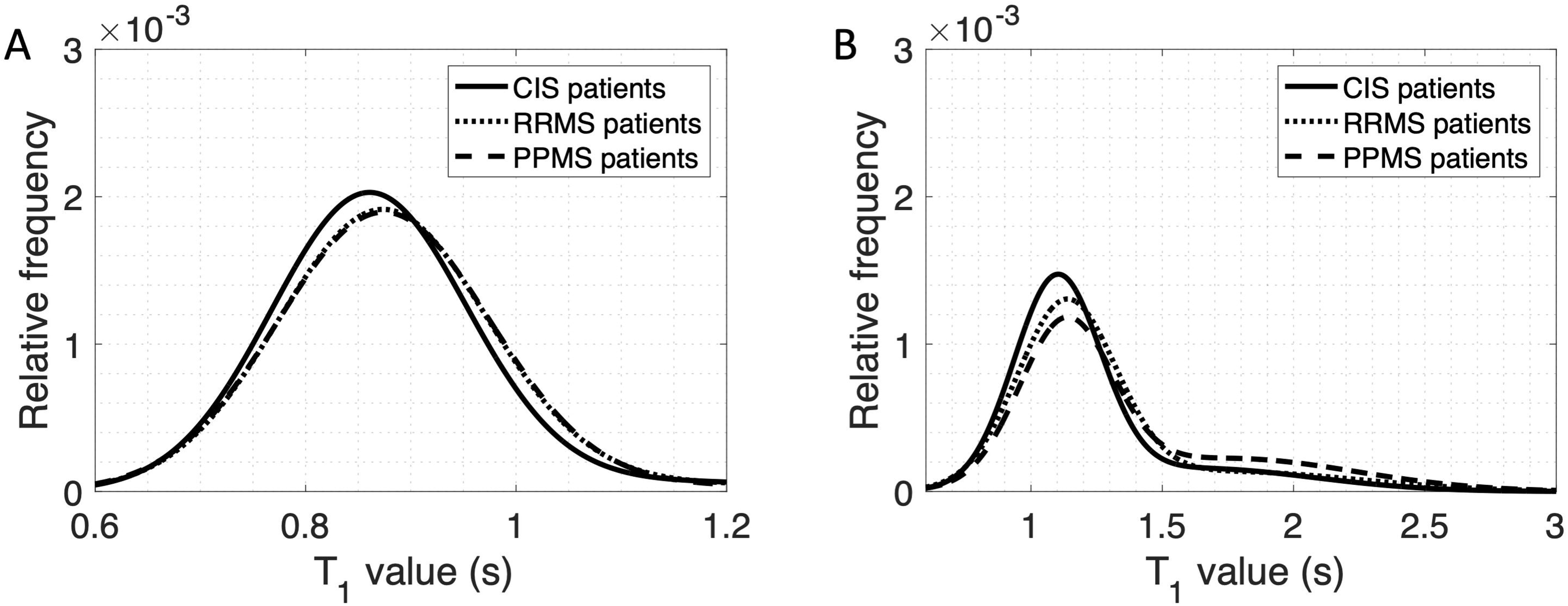
Distribution of T_1_ values in NAWM and MS lesions across different MS phenotypes. (A) T_1_ values measured in NAWM. (B) T_1_ values measured in MS lesions. Data are shown for patients with CIS, RRMS, and PPMS phenotypes. MS indicates Multiple Sclerosis; CIS indicates Clinically Isolated Syndrome; RRMS indicates Relapsing-Remitting Multiple Sclerosis; PPMS indicates Primary Progressive Multiple Sclerosis; NAWM indicates Normal-Appearing White Matter.

**Table 2.** Global analysis of T_1_ values in the NAWM of MS patients with different phenotypes and their corresponding control groups. For each group, the reported values correspond to the mean and standard deviation calculated from individual subject measurements. *p < 0.05 compared to corresponding controls; ***p < 0.001 compared to corresponding controls; ^||^p < 0.01 compared to PPMS patients; ^|||^p < 0.001 compared to PPMS patients. CIS indicates Clinically Isolated Syndrome; RRMS indicates Relapsing-Remitting Multiple Sclerosis; PPMS indicates Primary Progressive Multiple Sclerosis; NAWM indicates Normal-Appearing White Matter; FWHM indicates Full Width at Half Maximum.

| **Group type** | **CIS**<br>**(n=32)** | **Matched-**<br>**CIS-**<br>**controls**<br>**(n=43)** | **RRMS**<br>**(n=30)** | **Matched-**<br>**RRMS-**<br>**controls**<br>**(n=35)** | **PPMS**<br>**(n=25)** | **Matched-**<br>**PPMS-**<br>**controls**<br>**(n=26)** |
| --- | --- | --- | --- | --- | --- | --- |
| **Mean $T_1$**<br>**(s)** | 0.881 ±<br>0.017 | 0.885 ±<br>0.019 | 0.888 ±<br>0.019 | 0.884 ±<br>0.016 | 0.886 ±<br>0.021 | 0.894 ±<br>0.019 |
| **FWHM (s)** | 0.219 ±<br>0.010 | 0.214 ±<br>0.010 | 0.226 ±<br>0.013* | 0.218 ±<br>0.011 | 0.235 ±<br>0.009*** | 0.215 ±<br>0.008 |
| **Skewness** | 1.945 ±<br>0.173$^{\text{III}}$ | 1.933 ±<br>0.194 | 2.064 ±<br>0.261***$^{\text{II}}$ | 1.880 ±<br>0.019 | 2.246 ±<br>0.206*** | 1.997 ±<br>0.222 |

**Fig. 6.**
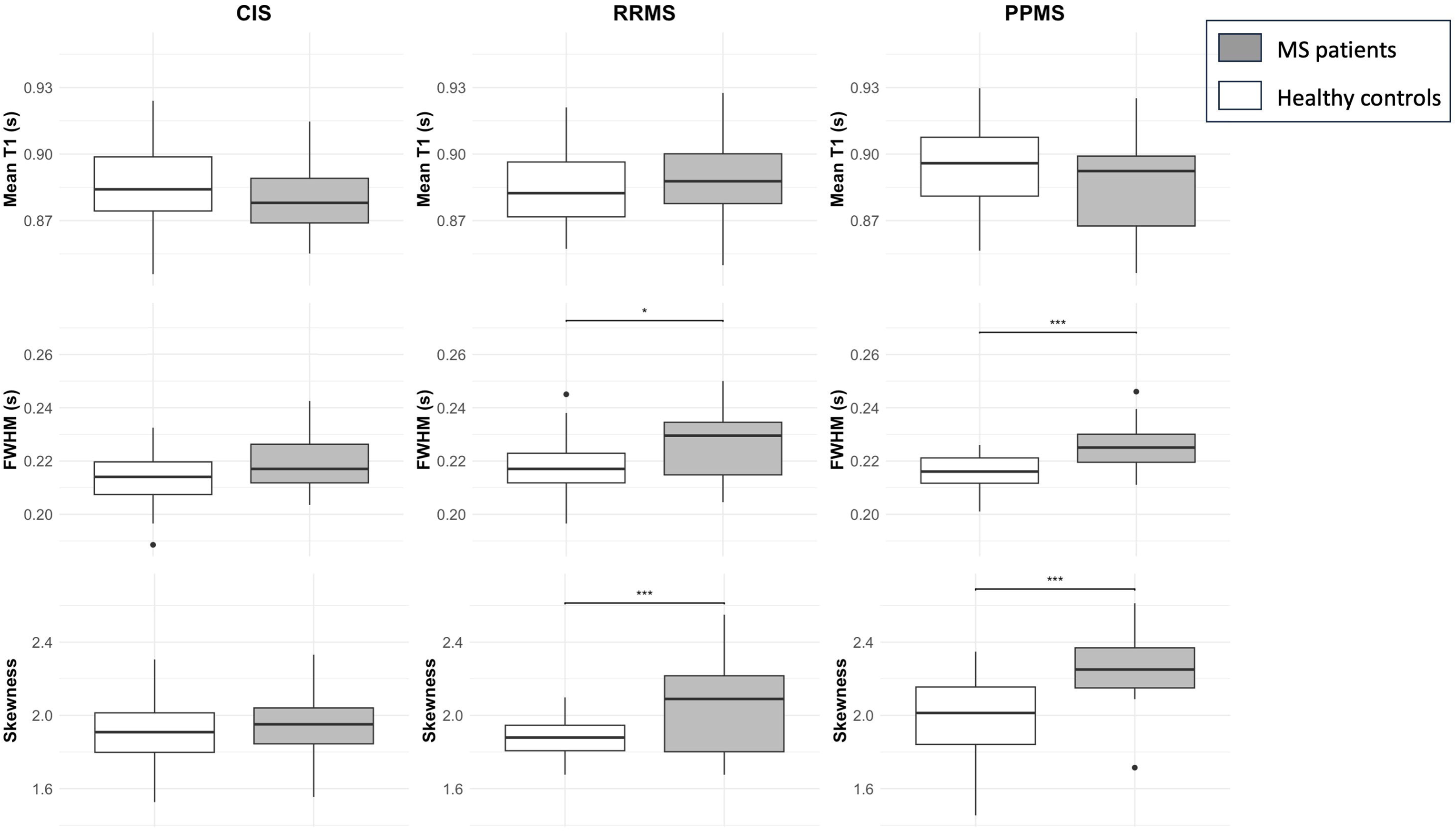
Comparison of T_1_ histogram metrics in the NAWM between patients (in gray) and matched controls (in white), for the three MS phenotypes: CIS, RRMS, and PPMS. The analyzed metrics include: mean T_1_, FWHM, and skewness. Boxes represent the median and interquartile range, and whiskers indicate data ranges. Boxes represent the median and interquartile range, whiskers extend to the data points within 1.5 x the interquartile range, and dots indicate individual outliers. Statistical significance between patients and controls is marked as follows: p < 0.05 (*), p < 0.001 (***). MS indicates Multiple Sclerosis; CIS indicates Clinically Isolated Syndrome; RRMS indicates Relapsing-Remitting Multiple Sclerosis; PPMS indicates Primary Progressive Multiple Sclerosis; NAWM indicates Normal-Appearing White Matter; FWHM indicates Full Width at Half Maximum.

In the CIS group, representing patients at an early disease stage, no statistically significant differences were observed between controls and patients for any of the measured parameters. In the RRMS and PPMS groups, while mean T_1_ values remained similar between patients and controls, differences in distribution were evident. These differences resulted in statistically significant variations in FWHM and skewness between MS patients and HC.

Regarding skewness, statistically significant differences were found between CIS and PPMS patients, as well as between RRMS and PPMS patients. However, no significant difference was observed between CIS and RRMS patients. Moreover, no statistically significant correlation was found between the histogram characteristics of NAWM and patient age in the MS cohort. Pearson correlation coefficients for the mean T_1_, FWHM, and skewness ranged from 0.063 to 0.19, with p-values between 0.09 and 0.54, suggesting no strong age effect on these parameters. Similarly, no significant correlation was observed between lesion volume and mean T_1_ values in the NAWM (r = 0.15; p = 0.16), indicating that lesion burden does not appear to be a predictive marker of the microstructural properties of the NAWM as measured by T_1_ value. An increase in T_1_ values was observed in lesions compared to NAWM (Fig. 5.B) across all patient groups. Specifically, the mean T_1_ values in lesions progressively increased from 1.219 ± 0.121 s in CIS patients, to 1.259 ± 0.084 s in RRMS patients, and 1.304 ± 0.109 s in PPMS patients, although no statistically significant differences were observed between groups.

## 4. Discussion

Quantitative T_1_ imaging is a key tool for assessing brain lesions in MS and, more broadly, in neurodegenerative diseases. While the MP2RAGE sequence is widely recognized as a reliable method for generating T_1_ maps in the brain, it remains underutilized in routine neuroradiology imaging protocols. In contrast, the MPRAGE sequence is routinely used in neuroradiology departments and is included in the vast majority of brain imaging protocols.

The objective of this study was to address the limited availability of dedicated T_1_ mapping acquisitions by leveraging standard MPRAGE scans. Specifically, we investigated the feasibility of generating clinically relevant quantitative T_1_ maps from MPRAGE acquisitions. As an initial step, we evaluated the approach in four healthy volunteers by comparing MPRAGE-derived T_1_ values obtained in various brain regions with those obtained with the MP2RAGE sequence. The results demonstrated a good correspondence of the two methods, particularly for T_1_ values below 1100 ms, which are typically attributed to WM. The T_1_ values obtained, apart from those from CSF, were also consistent with those reported in the literature (Bojorquez et al., 2017; Hagiwara et al., 2021), supporting the relevance of the approach.

For a proof-of-concept evaluation of the approach under disease conditions, this study specifically focused on detecting subtle abnormalities in NAWM across different MS phenotypes. Although these regions are not typically identified as lesional on conventional T_1_-and T_2_-weighted FLAIR and MPRAGE images, quantitative T_1_ values may provide early indicators of disease progression or treatment response.

The proposed T_1_ mapping method requires the definition of a reference ROI, for which the T_1_ value is known and fixed, in order to scale the correspondence curve between signal intensity and T_1_ values. In this study we choose the cingulate GM as the reference region for two main reasons. First, age-related variations in T_1_ are minimal in the gray matter, which allows the use of a single fixed T_1_ value for all subjects (Hagiwara et al., 2021). Second, among the gray matter regions, the cingulate cortex has been identified as exhibiting the smallest difference in T_1_ distribution between MS patients and HC (Steenwijk et al., 2016), despite potential variations in global NAGM T_1_ in MS patients (Bluestein et al., 2012; Gracien et al., 2016a; Vrenken et al., 2006). Indeed, Vrenken et al. reported T_1_ differences in NAGM between patients and controls, but the groups compared were not age-matched, which limits the ability to attribute the observed variations exclusively to pathology (Vrenken et al., 2006). Part of the observed differences may be related to age on top of MS-specific changes. Gracien et al. also reported significant T_1_ differences in NAGM at 3T, with relatively small variations on the order of 20 ms. However, these differences were not uniform across all NAGM regions, with some areas showing no significant changes (Gracien et al., 2016a). Finally, Bluestein et al. observed differences in global NAGM T_1_ at 7T between patients and controls, but measurements at high field strength are likely to amplify T_1_ variations, making the results difficult to compare to those of our 3T study (Bluestein et al., 2012).

In this study, we assumed that the cingulate GM was free of lesions, as lesions are particularly difficult to detect within the gray matter on conventional FLAIR or MPRAGE images. Future work could benefit from the use of double inversion recovery (DIR) sequences, which provide improved suppression of both CSF and WM signal and therefore enhance lesion visibility in the GM (Lecler, 2023; Lespagnol et al., 2023). Such an approach would allow a more reliable exclusion of possible lesions within the reference ROI and further strengthen the validity of the fixed T_1_ assumption in this region.

The T_1_ mapping method also accounts for proton density differences across tissues (Lavielle et al., 2023). Distinct values have been assigned to WM and GM proton densities for controls and MS patients, based on results from the literature (Gracien et al., 2016b; Hagiwara et al., 2021). For comparison, T_1_ maps were also generated under the assumption of identical proton density values in both groups. The difference between the T_1_ values obtained with the two different approaches was less than 0.2%, highlighting the minimal impact of proton density variations. This minimal difference arises from the nearly identical ratio of WM to GM proton density in patients and controls. The sensitivity of the method to errors in proton density estimation has been previously evaluated (Lavielle et al., 2023).

T_1_ measurements in the WM of 125 HC confirmed an age-related increase in T_1_ values, a finding consistent with previous literature (Hagiwara et al., 2021). Additionally, T_1_ values found in this study were within the expected range for WM at 3T (Bojorquez et al., 2017; Hagiwara et al., 2021), further reinforcing the validity of our T_1_ mapping approach.

Regarding MS patients, T_1_ values and distributions in the CIS group were highly similar between patients and controls. This similarity is consistent with CIS being an early stage of the disease. However, in RRMS and PPMS patients, an increase in T_1_ value was observed in both NAWM and MS lesions compared to CIS patients. These findings are in line with a more advanced stage of disease in RRMS and PPMS patients than in CIS patients. Despite similar mean T_1_ values between patients and controls in these groups, differences in distribution were evident, leading to statistically significant variations in FWHM and skewness, in accordance with previous findings (Parry et al., 2002; Steenwijk et al., 2016). These results are consistent with findings from other advanced MRI techniques. For instance, a reduced Magnetization Transfer Ratio has been observed in MS patients compared to HC (Filippi et al., 2000; Khaleeli et al., 2007; Tortorella et al., 2000). Similarly, Diffusion-Weighted Imaging has shown increased Mean Diffusivity and decreased Fractional Anisotropy values not only within focal lesions but also in the NAWM of MS patients (Bammer et al., 2000; Filippi et al., 2001). These findings reflect a diffuse tissue alteration extending beyond focal lesions, driven by a process of demyelination and axonal loss.

T_1_ values were consistently higher in MS lesions than in NAWM in all patient groups. In addition, T_1_ values of lesions were higher in RRMS and PPMS patients than in CIS patients. This variation in T_1_ values between clinical types probably reflects underlying differences in tissue composition and pathological processes (Brex et al., 2000). T_1_ measurements in lesions should be interpreted with caution, as the proton density used for T_1_ map computation was assumed to be equivalent to that of NAWM. However, proton density variations within MS lesions remain poorly characterized in the literature. Despite this assumption, our results demonstrate a clear trend of increasing T_1_ with disease severity. Applying a uniform proton density correction to all lesions would shift all T_1_ values in the same direction, preserving the observed trend.

## 5. Conclusion

The results confirm the feasibility and relevance of quantitative T_1_ measurements derived from an MPRAGE anatomical imaging protocol in MS patients. This proof-of-concept study demonstrates that with the proposed approach, it is possible to obtain reliable quantitative information, showing similar values to that provided by dedicated T_1_ mapping sequences, without requiring additional acquisitions. Given the widespread use of the MPRAGE sequence, this novel approach has the potential to significantly improve the quality of MS patient follow-up and treatment monitoring.

## Data Availability

All data produced in the present study are available upon reasonable request to the authors.

## Statements and declarations

### CRediT authorship contribution statement

**Audrey Lavielle**: Conceptualization, Data curation, Formal analysis, Software, Validation, Methodology, Visualization, Writing - original draft

**Fanny Munsch**: Resources, Validation, Writing - review & editing

**Aurélie Ruet**: Investigation, Resources, Validation, Writing - review & editing

**Thomas Tourdias**: Supervision, Validation, Writing - review & editing

**Yannick Crémillieux**: Conceptualization, Supervision, Resources, Project administration, Formal analysis, Validation, Methodology, Writing - review & editing

### Human rights

The authors declare that this work was performed in accordance with the Declaration of Helsinki of the World Medical Association revised in 2013 for experiments involving humans.

### Informed consent and patient details

The authors declare that this report does not contain any personal information that could lead to the identification of the patients.

